# Towards clinical implementation of artificial intelligence in cancer care: Concept mapping analysis of provincial workshop findings

**DOI:** 10.64898/2026.03.26.26349205

**Authors:** Hong Hao Xu, Chhavi Nayyar, Daniel Hilbers, Jonathan Avery, Srinivas Raman, Alan T. Bates, Cristina Conati, Ahmad Fayaz-Bakhsh, John-Jose Nunez

## Abstract

**Background:** Artificial intelligence (AI) has rapidly garnered interest in healthcare. Cancer care’s multidisciplinary nature and high coordination demands are well positioned to benefit from AI. While attitudes toward implementation of AI in medicine have been explored generally, literature remains scarce with specific regards to AI in cancer care. This study sought to understand perspectives of both patients and professionals in guiding responsible, effective implementation of evidence-based AI in cancer care.

**Methods:** We conducted a workshop at a provincial Cancer Summit (Vancouver, Canada). Discussions addressed concerns, benefits, and priorities for AI in cancer care. Responses from 48 workshop participants underwent structured conceptualization by concept mapping. Sorting and rating of the resulting statements were performed by a purposively sampled 13-member expert panel. Multidimensional scaling, hierarchical cluster and subcluster analysis produced visual and quantitative maps of findings.

**Results:** A total of 265 statements on perceived benefits, concerns, and priorities related to the implementation of AI in cancer care were generated; a deduplicated and consolidated final set of 100 statements underwent concept mapping. Two main clusters identified pertained to “Challenges and Safeguards for AI Implementation “ (Cluster 1) and “Clinical Benefits and Efficiency Gains” (Cluster 2). Subcluster analysis distinguished 8 thematic subclusters (4 per cluster). Mean importance and feasibility ratings were higher for Cluster 2, with large effect sizes for both importance (Cohen’s d = 0.94) and feasibility (Cohen’s d = 1.59). Ratings by clinical and nonclinical professionals were similar across comparisons except for Cluster 2 feasibility, rated higher by clinical participants (*P* = .029, Hedges’ g = 0.465). Further go-zone analysis classified statements according to their relative superiority/inferiority in importance and feasibility from overall average.

**Conclusions:** Expert panel ratings were higher for statements describing clinical benefits and efficiency gains than for those describing challenges and safeguards for AI implementation in cancer care. Concept mapping analysis distinguished between workflow-aligned AI applications, perceived as ready for implementation, and system-level governance requirements requiring longer-term investment. Present findings offer initial, context-specific structuring of stakeholder perspectives that may inform the prioritization and sequencing of AI implementation efforts in cancer care, providing a foundation for validation in other settings.

**Contributions to the literature:**

- While systematic reviews have identified barriers and facilitators to AI adoption in healthcare, this study provides the first structured prioritization framework for AI implementation in cancer care using concept mapping methodology, addressing an urgent need as AI capabilities advance rapidly in oncology.
- Strategic-level stakeholders distinguished between workflow-aligned domains perceived as implementation-ready and system-level domains requiring longer-term investment, enabling sequenced implementation planning that matches organizational readiness.
- This approach, combining broad stakeholder statement generation with expert panel prioritization, demonstrates how integrating experiential knowledge with quantitative prioritization methods can provide implementation scientists and health system leaders with actionable frameworks for staged AI adoption strategies.

## BACKGROUND

Artificial intelligence (AI) may be best understood as the emulation of human intelligence and cognitive processes by computer systems[1]. Recently, AI technologies have been undergoing rapid and substantial growth, sparking major enhancements across systems and processes of virtually all industries[2,3]. In healthcare, AI likewise holds a prominent and promising role that has the potential to drastically improve care[4,5]. Virtually every medical specialty, including specialties involved in cancer care[6], has been seeking to harness the benefits of AI in their practices, as seen by an ever-growing body of AI-related literature in recent times[7–10].

Understanding implementation of complex innovations like AI requires theoretical grounding. The Consolidated Framework for Implementation Research (CFIR) provides a comprehensive taxonomy of implementation determinants across five domains: intervention characteristics, outer setting, inner setting, characteristics of individuals, and implementation process[11]. Integrating CFIR with participatory methods like concept mapping enables both theory-driven identification and stakeholder-driven prioritization of implementation factors, making it particularly suited for translating framework constructs into actionable implementation strategies.

A key aspect in the implementation of new paradigms and translation of new evidence (e.g., evidence-based [EB] AI) is to evaluate stakeholders’ attitudes[12–14] and ascertain the acceptability and satisfaction[15–16] of the latest research findings about new services and technologies among all stakeholders including payers, providers and patients[17]. In the context of EB innovations and implementation, stakeholders are not limited to clinical end users: they also include the knowledge creators (i.e., AI engineers) and AI knowledge brokers (implementation scientists)[11,18]. Adequate attention to the attitudes of all stakeholders has been shown to facilitate collaboration[11,18]. Conciliatory attitudes towards EB innovation and its resulting stakeholder participation should be encouraged, and be continued throughout the entire stages of research and implementation: such as in co-identification of AI research agenda, collaboration in research conduct and co-creation of new AI knowledge, and finally in co-selection of implementation strategies and implementation outcome indicators[11]. Otherwise, mere awareness of the latest worldwide research findings on cancer or other disease risk factors[19–20] and treatments[21], or even knowledge of locally grown and hence relevant science, has historically not automatically translated to knowledge uptake and the consequent meaningful improvements in healthcare, health or human development[22,23]. At best, it has caused fractional uptake after considerable translation delays exceeding a decade[24,25]. Implementation science theories, particularly normalization process theory, emphasize that successful adoption depends not only on innovation characteristics but also on contextual factors and stakeholder engagement. Past research has demonstrated that conciliatory attitudes toward EB innovations, and hence improved evidence uptake, could occur through many means, including advanced participatory research powered by modern technologies[26], such as AI itself[27]. However, it is argued that collaboration in implementation research could also take place through the use of less fashionable methods, such as past research on the impact of asynchronous telemedicine to improve service demand by patients or to improve uptake by care suppliers of innovative computerized systems[21,28,29].

In this regard, scarce but otherwise valuable work has emerged which looks at patient and healthcare professionals’ attitudes towards AI in medicine at large[30–35]. Yet, there lacks a dedicated effort specific to oncology to account for the field’s unique intricacies, with one review study looking at patient attitudes toward AI in cancer care[36]. In this context, our group sought to conduct a participatory, collaborative and multi-step study of the attitudes of AI stakeholders, including AI engineers and developers as well as cancer patients and health professionals alike. In doing so, we focused on the implementation of AI tools and technologies in cancer care. Our project consisted of an interactive and collaborative workshop at a provincial cancer summit, followed by a mixed-method participatory data collection and analysis through concept mapping. The objective of the study was to identify which perceived benefits, concerns, and priorities matter most — and are most actionable — for the implementation of AI in cancer care, and to derive from these the practical implications for sequencing implementation efforts. The goal of the study was to inform research priorities and guide future work on the selection of best AI implementation strategies and AI implementation outcome measures according to the needs of the cancer care community.

## METHODS

### Theoretical Framework

This study was guided by the CFIR, which provides a comprehensive taxonomy for identifying and organizing implementation determinants. CFIR’s five domains (intervention characteristics, outer setting, inner setting, characteristics of individuals, and implementation process) served as our deductive framework for organizing stakeholder-generated statements. We integrated CFIR with concept mapping methodology, a participatory approach grounded in community-based participatory research principles, to ensure stakeholder perspectives shaped both identification and prioritization of implementation factors. This integration allows for theory-informed categorization while maintaining stakeholder-driven prioritization, addressing a key gap in implementation research: translating comprehensive frameworks into actionable, context-specific priorities.

### Study Context: BC Cancer Summit Workshop

The BC Cancer Summit is an annual oncology event in the Canadian province of British Columbia. It is hosted by BC Cancer, the province’s comprehensive cancer control program, providing education, professional development, and relationship-building opportunities for oncology professionals from all cancer disciplines, in addition to patients, family members, community providers, researchers, and members of non-profit organizations[37,38]. On November 21, 2024, our team held an in-person workshop at the Summit, entitled “How Do You Want Predictive Artificial Intelligence Tools to Be Deployed at BC Cancer? An Interactive Session.” The workshop addressed 3 guiding questions: (1) concerns regarding the integration of AI into cancer care, (2) perceived benefits of AI, and (3) priorities for AI implementation and research. The session combined a brief overview of relevant AI applications with small-group discussions, structured prioritization exercises, and a closing plenary to synthesize key themes.

### Participants

Conduction of study activities was approved by the University of British Columbia (UBC) Research Ethics Board (H24–03306). The workshop was attended by 48 participants who consented to the collection of data for research purposes; this was almost all participants, besides a few who were intermittently present. The participants included cancer patients, family members, AI and computer science researchers, cancer researchers, oncology specialists, family physicians, other allied health clinicians, information technology professionals, and individuals in administrative or leadership positions within the healthcare system. Because data collection was integrated into a live summit session, individual-level demographic information (e.g., age, gender, ethnicity, or patient versus professional status of each contributor) was not collected from workshop participants. For the purpose of this concept mapping study, workshop participants assumed the brainstorming part. As for the sorting and rating parts of concept mapping, an expert panel of 13 members was recruited through purposive sampling to ensure representation across key domains relevant to AI implementation in cancer care. Consistent with concept mapping methodology, statement generation and statement structuring are distinct methodological phases: broad, diverse participation is essential for generating a comprehensive statement pool, whereas the subsequent sorting and rating tasks serve to organize an already established pool and are appropriately performed by participants selected for topical expertise rather than statistical representativeness[39,41,42]. Panel members were therefore purposively selected for their familiarity with AI implementation in cancer care, and panels of approximately 10–20 sorters have been shown to produce stable concept maps[39]. All panel members were affiliated with academic or healthcare institutions, stemming from various and diverse backgrounds spanning clinical cancer patient care, implementation science, computer science, and health system administration. All participants (n = 13, 100%) were involved in cancer research. Of these, 31% (n = 4) had specific expertise in AI research and development, 54% (n = 7) were clinicians, 8% (n = 1) were health administrators and patient-experts each. The clinical participants had backgrounds spanning medical oncology, radiation oncology, primary care, nursing, cancer psychiatry, and cancer dietetics, ensuring comprehensive representation across cancer care specialties. All 13 experts provided valid sorts and ratings of importance and feasibility of concept mapping statements. For subgroup analysis purposes, participants were categorized as clinical (n = 7, 54%) or non-clinical (n = 6, 46%) based on their primary role. Clinical participants included those with direct patient care responsibilities across various cancer care specialties.

### Concept Mapping Procedure

Concept mapping is a structured participatory method grounded in community-based participatory research principles, which posit that those affected by implementation should be central to identifying and prioritizing barriers and facilitators. It has been widely applied in healthcare and implementation research to organize stakeholder perspectives and inform planning[39–45]. In this study, concept mapping was performed following Trochim’s sequential 6-stage framework: (1) preparation, (2) generation, (3) organization, (4) representation, (5) interpretation, and (6) utilization[41].

The first 2 stages, preparation and generation, were accomplished through the workshop, where statements were generated by participants. Statement organization (stage 3) was performed independently by two authors (H.H.X and D.H.): raw statements were minimally edited for grammatical clarity while preserving meaning, deduplicated, and consolidated into a final 100-statement set via discussion to consensus, then shuffled from their original order using random number generation. The 13-member panel performed sorting and rating on Qualtrics XM (Qualtrics LLC, Provo, UT, USA). Sorting involved grouping all statements into arbitrary groups “in a way that makes sense to [the person performing the sorting task]”[42]. Rating used a 5-point Likert scale for (1) importance and (2) feasibility[42]. Importance was rated on a scale from 1 (relatively unimportant) to 5 (extremely important), relative to other statements. Because the statement pool comprised three types of statements (benefits, concerns, and priorities), raters were provided with a single, action-oriented definition of feasibility to be applied consistently across all statement types: feasibility was defined in the rating instructions as how easy it will be to implement a priority, actualize a benefit, or address a concern, rated on a scale from 1 (relatively unfeasible) to 5 (extremely feasible), relative to other statements. Under this instruction, rating a concern for feasibility referred to the ease of addressing or mitigating that concern, rather than to the likelihood of the concern occurring. This definition was displayed to raters within the Qualtrics rating task. All operations, including statement processing and panel selection, were performed following established concept mapping documentation and literature to ensure validity[39,41,42,46–48].

### Analysis

Importance and feasibility ratings were verified for completeness and organized into matrices for analysis. Multidimensional scaling (MDS) was applied to generate a two-dimensional representation of the statements. A co-occurrence matrix was constructed from participant sorting data by counting how frequently each pair of statements was sorted into the same pile across all participants, normalized by the number of participants. This co-occurrence matrix was converted to a distance matrix, and MDS was applied using a precomputed dissimilarity matrix with a random seed for reproducibility. This approach follows established concept mapping methodology where statement proximity reflects how often participants grouped statements together during the sorting phase[41]. Stress values were computed to evaluate the goodness-of-fit, with lower values indicating a more accurate representation of the original high-dimensional distances.

Hierarchical cluster analysis was performed on the MDS coordinates using Ward’s method, selected for its ability to produce compact, interpretable clusters suitable for concept mapping’s clear conceptual groupings. The optimal number of clusters was determined using silhouette analysis and the elbow method to assess cluster cohesion and separation. Consistent with literature recommendations, clusters were labelled through a conceptual interpretive approach utilizing iterative review of statement content, mean ratings, and thematic density[49].

A secondary hierarchical cluster analysis was performed to identify subclusters within main clusters. This allows for further fine-grained conceptual distinctions. Subclustering was performed separately within each main cluster using Ward’s method on the MDS coordinates of statements belonging to that cluster. The optimal number of subclusters was determined through visual inspection of dendrograms and consideration of conceptual coherence, resulting in 4 subclusters within each main cluster. Subclusters were labeled using the same interpretive approach and according to their position within the hierarchical structure (e.g., Subcluster 1.1, 1.2, 1.3, 1.4 within Cluster 1, and Subcluster 2.1, 2.2, 2.3, 2.4 within Cluster 2).

Likert ratings of importance and feasibility were analyzed at the statement, cluster, and subcluster levels. For individual statements, mean importance and feasibility scores, standard deviations, and response counts were calculated, and gap analyses were performed by subtracting feasibility from importance ratings. At the cluster level, aggregated means were computed, clusters were compared, and strategic priorities were identified based on observed patterns in importance and feasibility. At the subcluster level, mean importance and feasibility ratings were calculated, enabling more nuanced comparisons of strategic priorities within the broader cluster structure. Pattern match analysis was conducted to examine the relationship between importance and feasibility ratings across subclusters, revealing distinct patterns that inform implementation strategies. Finally, we conducted subgroup analysis, comparing importance and feasibility ratings between clinical and nonclinical participants at both the cluster and subcluster levels. Comparative analyses were performed using the Mann–Whitney U test and Student’s t-test with Cohen’s d for effect size. Given that ratings were nested within a small number of raters (n = 13) and statements do not constitute fully independent observations, these comparisons should be regarded as exploratory and descriptive rather than confirmatory; accordingly, we emphasize effect sizes and the direction and consistency of observed patterns over *P* values, which are reported for completeness.

Strategic prioritization was performed using a go-zone analysis framework, which maps subclusters into 4 quadrants based on their mean importance and feasibility ratings relative to overall means. This approach enables identification of: (1) priority subclusters (high importance, high feasibility) representing immediate implementation opportunities; (2) strategic subclusters (high importance, low feasibility) requiring research and development investment; (3) quick wins (low importance, high feasibility) suitable for rapid implementation; and (4) low priority subclusters (low importance, low feasibility) that may be deferred. The subcluster-level go-zone analysis provides more actionable insights than statement-level analysis by grouping related concepts and reducing complexity while maintaining granularity.

The analysis was implemented in Python (version 3.12, Python Software Foundation, Wilmington, DE), adapting the concept mapping methodology from Bar & Mentch[49] for Python implementation. The code and scripts used for this analysis are publicly available on GitHub (https://github.com/chhaviiiii/concept_mapping).

## RESULTS

### Workshop Outcomes

Following the 2-hour workshop, 48 participants together contributed to generating a total of 265 statements on the implementation of AI in cancer care. Of these, 87 statements (32.8%) pertained to concerns, 72 (27.2%) pertained to perceived benefits, and 106 (40.0%) pertained to research priority, with respect to the 3 workshop discussion questions.

### Concept Mapping – Statements

Following statement processing, a final list of 100 statements was obtained and sorted and rated by the 13-member expert panel. The panelists sorted the card statements into an average of 5 piles (mean = 5.0; SD = 2.4; range = 2–9). Panelists took a mean of 42.8 minutes (SD = 19.8; range = 14.6–71.7) to complete both tasks. **Table 1** displays the final list of statements and their associated mean importance and feasibility ratings.

**Table 1.** Concerns, Perceived Benefits, and Implementation/Research Priorities Regarding AI in Cancer Care, with Mean Importance and Feasibility Ratings. Given that this table was larger than one A4 or Letter page in length, it was placed at the end of the document text file. It should be added after the first paragraph of the section entitled “Concept Mapping – Statements.”)

| # | Statement | Importance | Feasibility |
| --- | --- | --- | --- |
| 1 | Human oversight during implementation in early days | 4.25 | 3.42 |
| 2 | Concerns with confidentiality | 3.75 | 3.25 |
| 3 | Concerns with patient acceptability | 4.08 | 3.17 |
| 4 | Efficiency improved for documentation | 3.58 | 4.17 |
| 5 | False confidence | 3.33 | 2.73 |
| 6 | AI could provide alternate perspectives | 3.17 | 3.17 |
| 7 | AI could perpetuate existing biases | 3.33 | 3.25 |
| 8 | Minimizing bias and increasing performance and accuracy | 4.00 | 3.33 |
| 9 | Ensuring accountability and oversight of AI | 4.00 | 3.58 |
| 10 | Concerns with digital literacy and digital divide barriers | 3.08 | 2.75 |
| 11 | Better detection (of moles, cancer lumps) | 3.58 | 3.75 |
| 12 | Diversification of AI training data and sources | 3.33 | 3.33 |
| 13 | Too much bureaucracy slowing progress | 3.00 | 3.00 |
| 14 | Concerns with liability | 3.33 | 2.83 |
| 15 | AI could decrease false positives | 3.42 | 3.42 |
| 16 | Proactive preventative care | 3.17 | 3.00 |
| 17 | Benefits in equitable access | 3.33 | 2.92 |
| 18 | AI could free up time for pt care | 3.75 | 3.67 |
| 19 | Developing a rigorous privacy and quality assurance framework | 3.50 | 3.42 |
| 20 | AI could be misused | 3.17 | 2.75 |
| 21 | Helps with generation of ideas can act as a sounding board | 2.83 | 3.58 |
| 22 | Benefits with accessibility & communication (translating languages lay terms) | 3.25 | 3.58 |
| 23 | AI allows access vast information very quickly to help diagnosis | 3.75 | 4.00 |
| 24 | Benefits in early distress screening | 3.33 | 3.25 |
| 25 | Establishing proper consent | 3.00 | 3.08 |
| 26 | Concerns with job security | 2.42 | 2.25 |
| 27 | Benefits with patient access to accurate information | 3.25 | 3.55 |
| 28 | Concerns about personalizing care | 2.83 | 3.25 |
| 29 | Data governance policies and regulations to enhance data quality and accountability | 2.92 | 2.58 |
| 30 | Savings in time, can integrate different information and improve in treatment especially when there is less number of human specialist | 3.83 | 3.67 |
| 31 | AI could remove human bias | 3.17 | 2.75 |
| 32 | Regulation framework and a good understanding of the medico–legal implications | 2.83 | 2.25 |
| 33 | AI could lead to excessive human delegation, assuming that AI will take care of it! | 2.58 | 2.50 |
| 34 | Knowledge repositing for learning and research | 2.92 | 3.50 |
| 35 | AI could be of concern regarding its ongoing validation | 3.00 | 3.09 |
| 36 | AI could be of concern to patient privacy | 3.08 | 3.08 |
| 37 | Consultation recording and transcription | 3.17 | 4.18 |
| 38 | Ensuring cybersecurity of AI tools | 3.75 | 3.17 |
| 39 | Educating end-users about properly navigating AI | 3.83 | 3.58 |
| 40 | Concerns with protection of personal health identifiers | 3.42 | 3.00 |
| 41 | Facilitation of clinical research | 3.08 | 3.42 |
| 42 | Improve access to services | 3.67 | 3.58 |
| 43 | Ensuring accessibility and translation | 3.50 | 3.67 |
| 44 | AI could help improve accuracy | 3.58 | 3.75 |
| 45 | Embed research / quality improvement into all areas | 2.92 | 3.25 |
| 46 | Garbage in, garbage out' – data that the AI model is trained on must be good, clean, large, and diverse | 3.58 | 2.83 |
| 47 | Concerns with attrition of skills in healthcare providers | 2.58 | 2.58 |
| 48 | Preservation of human touch | 3.08 | 2.83 |
| 49 | Loss of human connection/interactions | 3.33 | 2.75 |
| 50 | Summarization of large amounts of text or data – increases digestibility/highlighting key points | 3.67 | 4.33 |
| 51 | Collate large databases | 3.25 | 4.17 |
| 52 | AI could engender issues with trust | 2.75 | 2.67 |
| 53 | Pooling large volume of data can improve accuracy of AI system and allow for more timely diagnosis and treatment recommendation | 3.58 | 3.75 |
| 54 | Concerns with responsibility accountability of AI recommendations | 3.17 | 3.00 |
| 55 | AI hallucinations | 3.17 | 2.83 |
| 56 | Multidisciplinary collaboration and training in developing AI models | 3.17 | 3.25 |
| 57 | 24/7 availability, AI doesn't get tired or brain fog | 3.42 | 4.25 |
| 58 | Noticing patterns and new things humans haven't noticed | 3.58 | 3.50 |
| 59 | Prediction/prognosis/treatment recommendations | 3.92 | 3.58 |
| 60 | Concerns with standards for quality, accuracy, etc. | 3.08 | 2.92 |
| 61 | Concerns with power outages and downtime procedures | 2.17 | 2.58 |
| 62 | Increased opportunities for innovation | 2.92 | 3.58 |
| 63 | Distress is nuanced and often detected in the unsaid of human communication and in a trusting therapeutic relationship | 3.42 | 2.50 |
| 64 | Personalization of AI support tools for patients | 3.58 | 3.33 |
| 65 | Concerns with difficulty to know older data for training and missing new breakthrough information | 2.92 | 2.92 |
| 66 | Concerns with consent of patient | 3.25 | 3.17 |
| 67 | Racism bias through research (history of medical research) | 3.08 | 2.50 |
| 68 | Phased roll-out: Start with simple, "low stakes" tasks continue in stages | 3.17 | 4.08 |
| 69 | Reduce admin burden | 3.92 | 4.17 |
| 70 | Potential in navigation and directing to self-management resources | 3.42 | 3.67 |
| 71 | Over reliance by physicians and reduced problem solving skills | 2.67 | 2.75 |
| 72 | Ability to make text/conversation more digestible for patients (less technical) | 3.50 | 3.83 |
| 73 | Ensuring clinical validation of AI tools | 3.50 | 3.42 |
| 74 | Concerns with maintaining up to date information, sources and programming | 2.83 | 3.42 |
| 75 | Data privacy concerns | 3.33 | 2.83 |
| 76 | Triage diagnostic results for clinicians | 3.67 | 3.67 |
| 77 | Concerns with going too far and can't come back | 2.33 | 2.50 |
| 78 | Control of data by corporations | 2.75 | 2.50 |
| 79 | AI could help address the human health resources issue | 3.58 | 3.25 |
| 80 | Faster assessments to inform decision-making | 3.50 | 3.58 |
| 81 | Concerns with transparency | 3.08 | 2.75 |
| 82 | AI could make mistakes | 3.33 | 3.00 |
| 83 | Concerns with stability of the system (during virus attack, power outage, system interruption) | 2.75 | 2.42 |
| 84 | Live translation and/or transcription of conversations | 3.73 | 4.08 |
| 85 | Establishing guidelines for "best practices" for training AI + clinical validation | 3.50 | 3.17 |
| 86 | Lack of training/knowledge for those using the AI tool in the real world | 3.17 | 3.25 |
| 87 | Can existing healthcare servers support the use of AI? | 2.17 | 3.08 |
| 88 | Ensuring interpretability of AI tools | 3.42 | 3.33 |
| 89 | Allow clinicians to focus on tasks requiring their expertise | 4.17 | 4.00 |
| 90 | Identifying and focusing on high-priority areas, such as prevention | 3.08 | 3.42 |
| 91 | Concerns that data is held by private companies | 2.92 | 2.42 |
| 92 | Maintaining competency with changing best practices and approved standards | 3.17 | 2.83 |
| 93 | Minority groups are being marginalized | 3.00 | 2.92 |
| 94 | Patient/parent/caregiver/provider acceptability | 3.92 | 2.92 |
| 95 | Data ownership and standards and guidelines on AI developed are not been established | 2.83 | 2.92 |
| 96 | Possibility of data breaches / data leaks | 3.00 | 2.83 |
| 97 | Potential for widening differential diagnosis, widening perspective of patient | 3.00 | 3.33 |
| 98 | Lists possible outcome and risks for the patient | 3.83 | 3.58 |
| 99 | Cybersecurity concerns | 2.75 | 2.42 |
| 100 | Automation of routine tasks | 4.00 | 4.08 |

### Concept Mapping – Visual Outputs

Results from statement sorting were subject to nonmetric MDS analysis, yielding a spatial representation of statements where statements grouped together the most often are located more proximately, while those grouped together less frequently are further apart, as shown in **Figure 1a**. The statements are numbered to aid in cross-referencing the spatial relationships of the points on the map with their labels enumerated in **Table 1**. Two statistically optimal clusters were identified using Ward’s hierarchical clustering, validated through silhouette analysis, elbow method, and gap statistic evaluation (**Figure 1b**). Cluster 1 contained 62 statements (62%) and Cluster 2 contained 38 statements (38%). The cluster boundaries showed clear spatial separation and internal cohesion, with statements within each cluster located proximally in the MDS space. Cluster 1 was labelled as “Challenges and Safeguards for AI Implementation,” whereas Cluster 2 was labelled as “Clinical Benefits and Efficiency Gains.”

**Figure 1.**
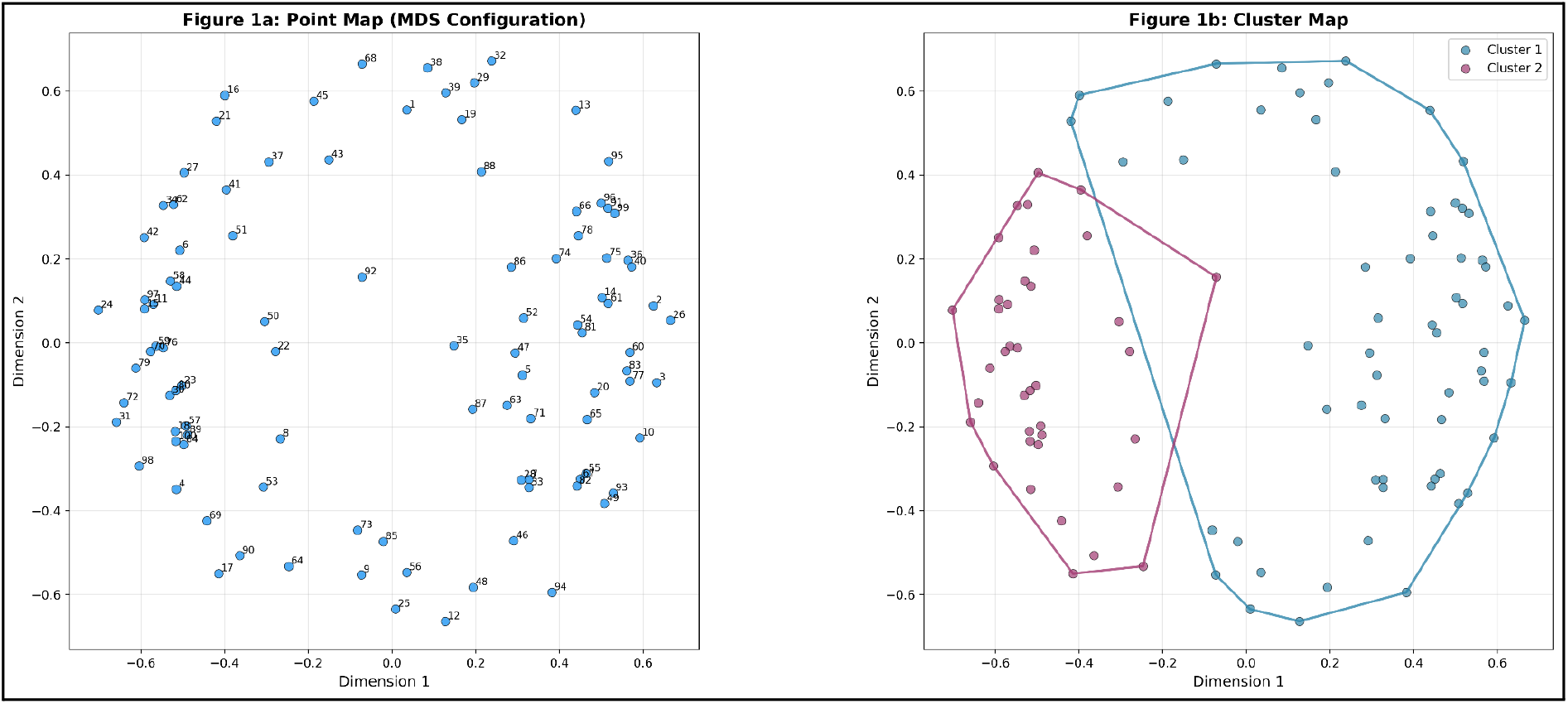
Point map that visually represents the relationships among the 100 statements, with each point on the map representing a statement. Figure 1b. Cluster map showing the final cluster solution with convex hull boundaries around each cluster.

The aggregated mean importance within each cluster was computed, with Cluster 2 rated higher (M = 3.50, SD = 0.32) than Cluster 1 (M = 3.14, SD = 0.43), corresponding to a large effect size (Cohen’s d = 0.94; exploratory t-test and Mann–Whitney U test both *P* < .001). Regarding feasibility, Cluster 2 was likewise rated higher (M = 3.62, SD = 0.38) than Cluster 1 (M = 2.99, SD = 0.41), again with a large effect size (Cohen’s d = 1.59; both tests *P* < .001). Given the small rating panel and non-independence of statements, these comparisons are interpreted as descriptive of the pattern within this panel rather than as confirmatory inference.

Secondary hierarchical cluster analysis identified subclusters within each main cluster, shown in **Figure 2**. Subclustering within Cluster 1 had 4 subclusters: Subcluster 1.1 (13 statements), Subcluster 1.2 (22 statements), Subcluster 1.3 (18 statements), and Subcluster 1.4 (9 statements). Cluster 2 also had 4 subclusters: Subcluster 2.1 (13 statements), Subcluster 2.2 (16 statements), Subcluster 2.3 (6 statements), and Subcluster 2.4 (3 statements). A full listing of statements assigned to each subcluster, including mean importance and feasibility ratings, is provided in **Additional file 1**. **Figure 2** further elucidates the finer-grained conceptual groups that maintain their position within the broader MDS space, revealing both the hierarchical structure and the nuanced relationships between related concepts.

**Figure 2.**
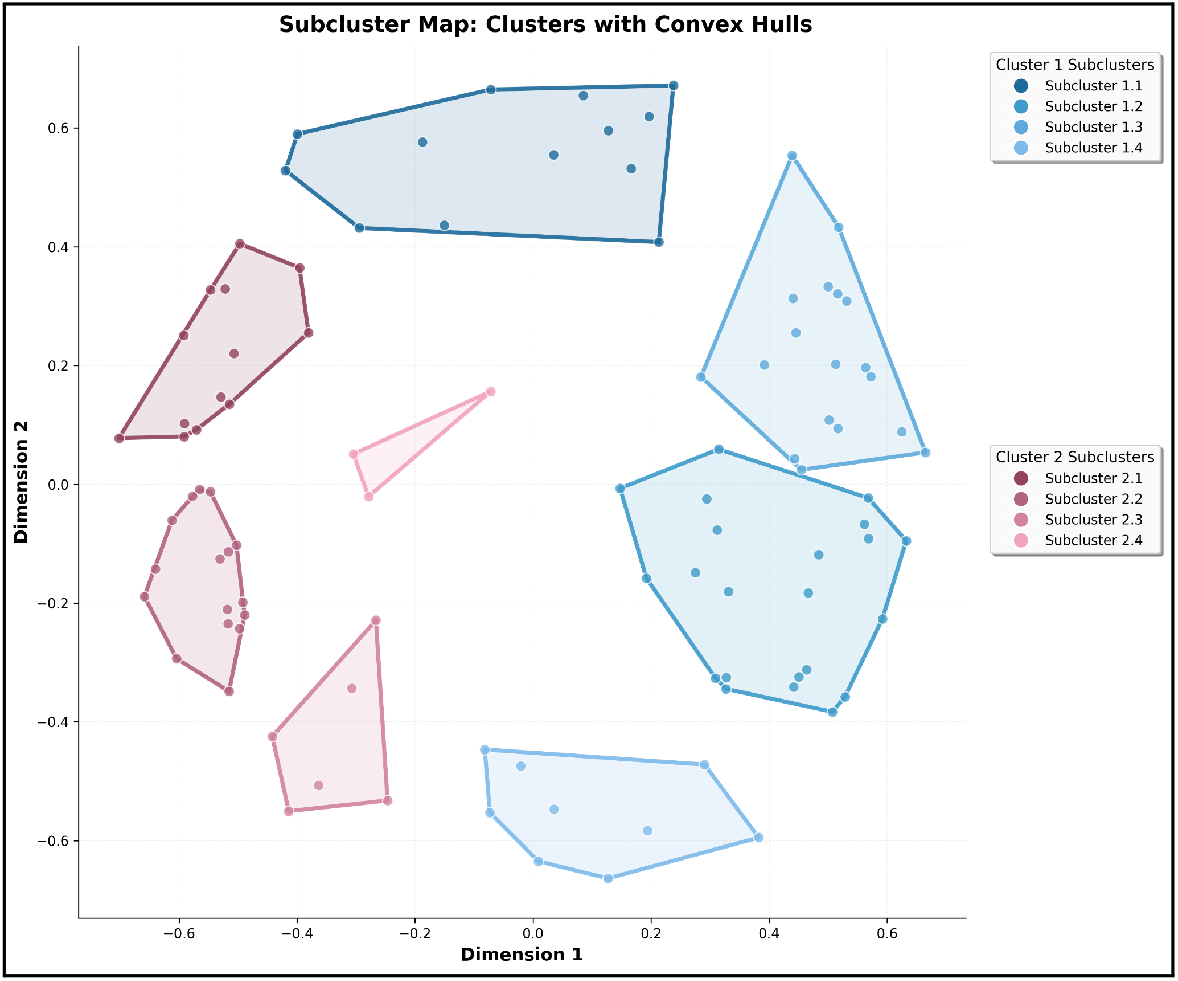
Subclusters within the MDS space. Subclusters 1.1–1.4 belong to Cluster 1 (Challenges and Safeguards for AI Implementation), and subclusters 2.1–2.4 belong to Cluster 2 (Clinical Benefits and Efficiency Gains)

The pattern match analysis (**Figure 3**) shows the relationship between importance and feasibility ratings at both cluster and subcluster levels. **Figure 3a** presents the cluster-level analysis, revealing distinct patterns in how the 2 clusters differ across rating dimensions. Cluster 2 demonstrates higher ratings in both importance (3.50) and feasibility (3.62), while Cluster 1 shows lower ratings in both dimensions (3.14 importance, 2.99 feasibility). Exploratory comparison using the Wilcoxon signed-rank test suggested differing importance–feasibility patterns within the two clusters: in Cluster 1, importance ratings tended to exceed feasibility ratings (*P* = .005), whereas in Cluster 2, feasibility ratings tended to exceed importance ratings (*P* = .007).

**Figure 3a.**
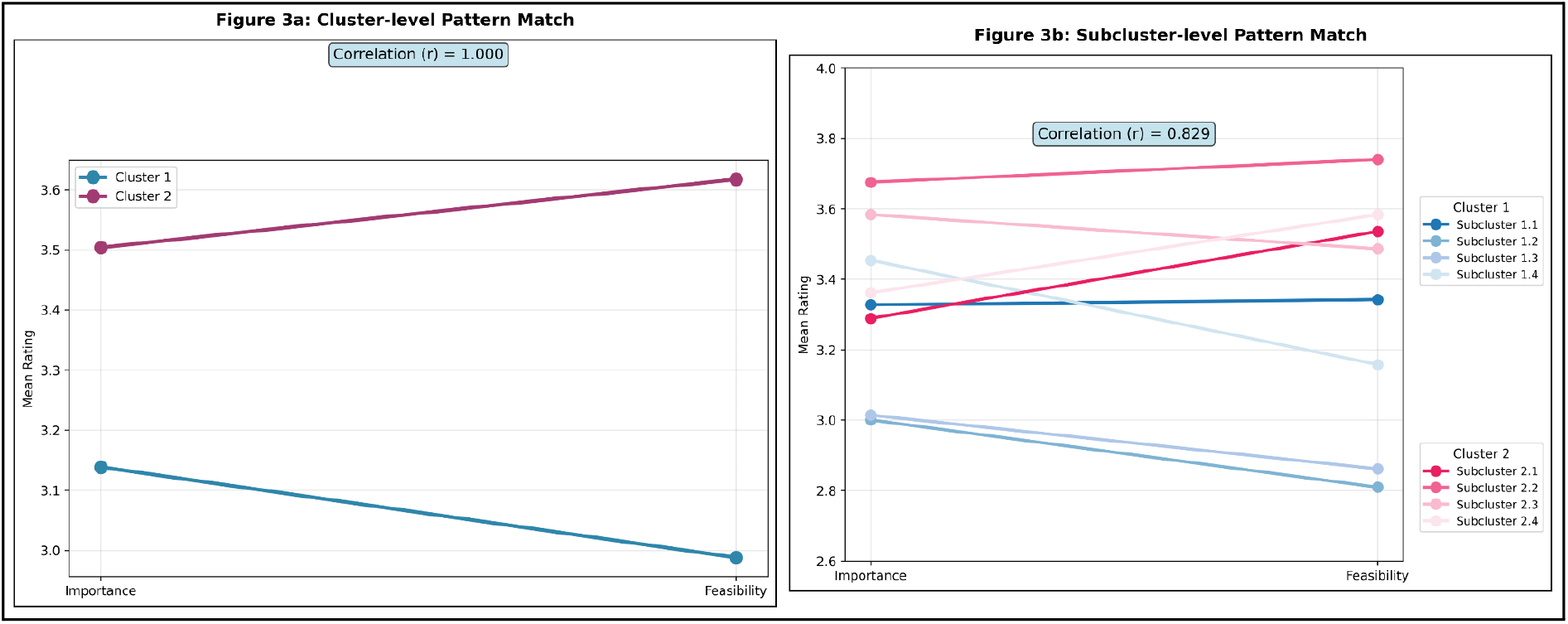
Cluster-level ratings, revealing distinct patterns between the two main clusters. Figure 3b. Analysis at the subcluster level, showing relationships across the eight subclusters with a correlation coefficient (r = 0.829) indicating a strong positive relationship between importance and feasibility across all subclusters.

**Figure 3b** extends this analysis to the subcluster level, revealing distinct patterns across the eight subclusters. Each subcluster is represented by a line connecting its mean importance rating to its mean feasibility rating, allowing for visual comparison of the gap between these dimensions. The analysis revealed a strong positive correlation between importance and feasibility ratings across subclusters (r = 0.829). However, notable variations exist in the magnitude of the gap between importance and feasibility across subclusters. Subclusters within Cluster 2 (Clinical Benefits and Efficiency Gains) generally demonstrate higher ratings in both dimensions compared to those in Cluster 1 (Challenges and Safeguards). Specifically, Subcluster 2.2 shows the highest mean ratings in both importance (3.68) and feasibility (3.74). This subcluster comprises the direct workflow-improvement statements (e.g., documentation efficiency, triage of diagnostic results, live translation and transcription, automation of routine tasks), while Subclusters 1.2 and 1.3 show the lowest ratings in both dimensions. The pattern match visualization highlights subclusters with differing relationships between importance and feasibility ratings.

Results from subgroup analysis comparing ratings of clinical and nonclinical panel members are shown in **Figure 4**. For Cluster 1, no difference was found for both importance (Clinical: 3.15 vs Non-Clinical: 2.97; *P* = 0.079) and feasibility (Clinical: 3.00 vs Non-Clinical: 2.84; *P* = 0.144) ratings. For Cluster 2, clinical participants provided somewhat higher mean feasibility ratings (Clinical: 3.64 vs Non-Clinical: 3.34; *P* = 0.029), though the effect size was small to medium (Hedges’ g = 0.465) and this exploratory comparison was based on very small subgroups. Mean importance ratings did not differ appreciably (Clinical: 3.52 vs Non-Clinical: 3.32; *P* = 0.128).

**Figure 4.**
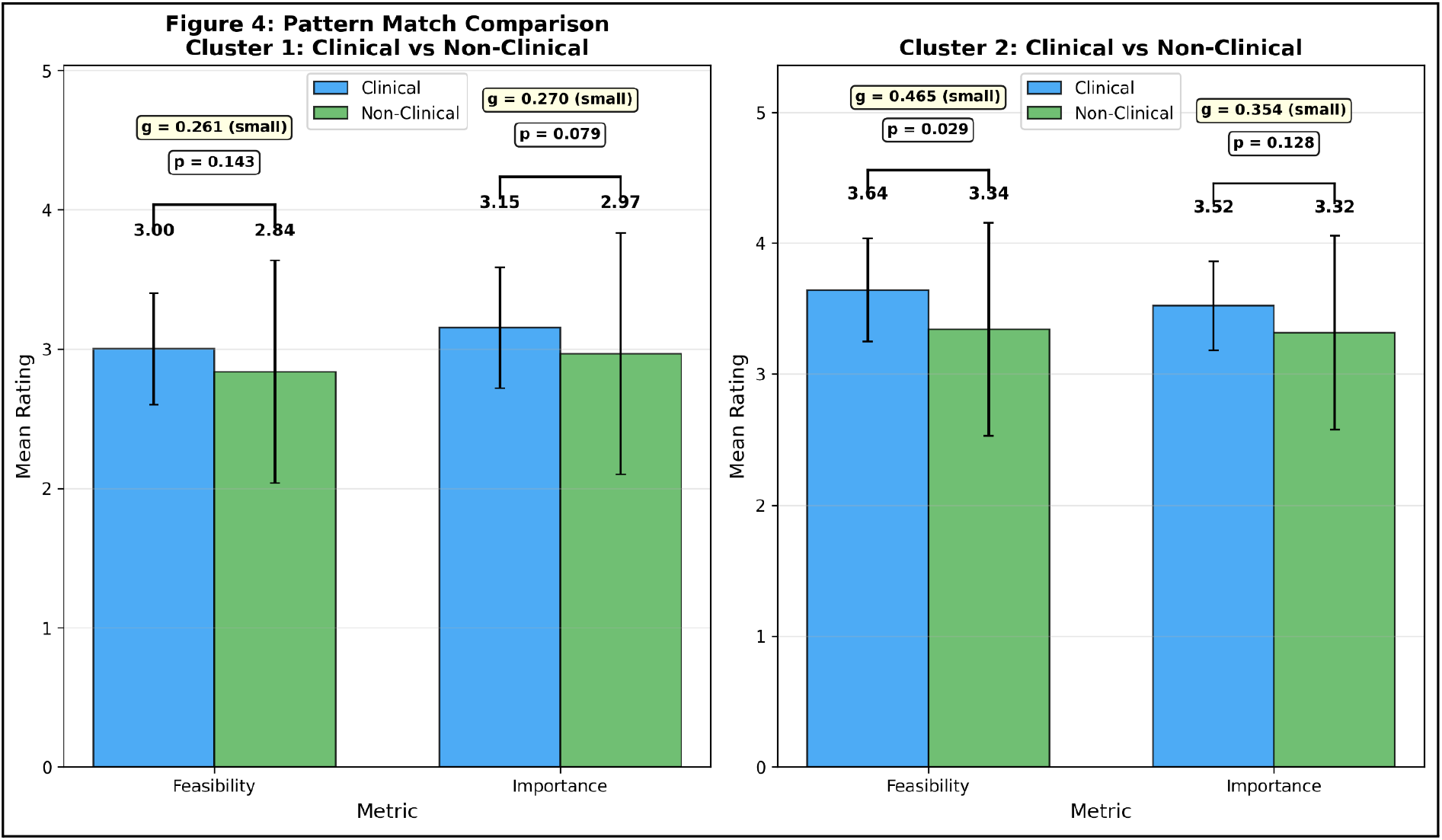
Subgroup analysis of mean importance and feasibility ratings between clinical and nonclinical professionals.

Finally, the go-zone plot is shown in **Figure 5**, which positions statements relative to the average importance and feasibility ratings. The plot is divided into four quadrants based on overall mean values for each rating scale. Quadrant I (High–High; 36 statements, 36%), representing immediate implementation opportunities, was dominated by workflow-embedded applications: the highest-rated statements included allowing clinicians to focus on tasks requiring their expertise (importance 4.17, feasibility 4.00), automation of routine tasks (4.00, 4.08), reducing administrative burden (3.92, 4.17), summarization of large amounts of text or data (3.67, 4.33), and documentation efficiency (3.58, 4.17). Notably, this quadrant also contained the single highest-importance statement overall: human oversight during early implementation (4.25, 3.42), alongside accountability and oversight of AI (4.00, 3.58), indicating that the panel viewed certain governance functions as both critical and attainable. Quadrant II (Low–High; 16 statements, 16%) comprised quick wins such as collating large databases (3.25, 4.17) and consultation recording and transcription (3.17, 4.18). Quadrant III (High–Low; 13 statements, 13%), representing research and development priorities, was populated chiefly by acceptability and trust-related requirements: patient acceptability (4.08, 3.17), patient/parent/caregiver/provider acceptability (3.92, 2.92), cybersecurity of AI tools (3.75, 3.17), training data quality (“garbage in, garbage out”; 3.58, 2.83), and the difficulty of detecting nuanced distress within a trusting therapeutic relationship (3.42, 2.50). Quadrant IV (Low–Low; 35 statements, 35%) contained most system-level governance and risk statements rated as lower priority, including regulatory frameworks and medico-legal implications (2.83, 2.25), corporate control of data (2.75, 2.50), and job security concerns (2.42, 2.25). A per-quadrant compilation of statements from the go-zone plot is further provided in **Additional file 2.**

**Figure 5.**
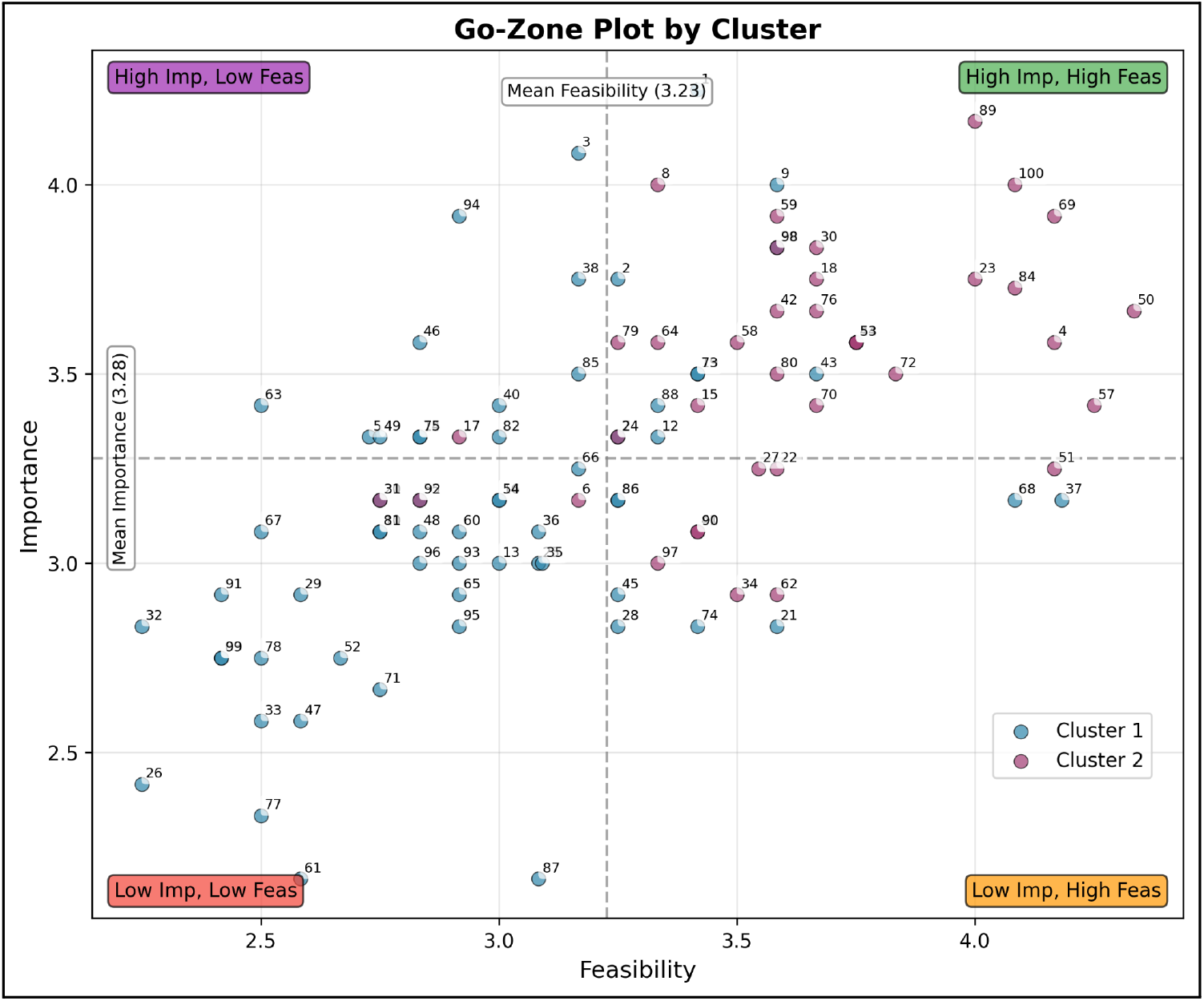
presents the go-zone plot identifying statements based on their performance relative to average importance and feasibility ratings.

## DISCUSSION

To our knowledge, this study represents the first use of concept mapping to examine end-user perspectives on AI implementation in cancer care. Prior work, including systematic reviews, has synthesized barriers and facilitators thematically; however, these studies have not applied a structured, participatory approach that organizes stakeholder-generated statements into relational clusters and quantitatively derived priority spaces. Concept mapping served here as the means to a substantive end: identifying which implementation considerations informed participants judged most important and most actionable in cancer care, and what those judgments imply for how organizations sequence implementation efforts. This approach aligns with recent concept mapping studies that emphasize the importance of combining stakeholder engagement with quantitative structuring to inform implementation planning and strategy selection[44,45].

In interpreting these results, it is important to distinguish the two participatory phases of this study. The statement pool reflects the perspectives of a broad, multi-stakeholder workshop audience (n = 48), whereas the priority structure, the clusters, ratings, and go-zone placements, reflects the judgments of a smaller, predominantly professional expert panel (n = 13, including one patient-expert). The “priorities” reported here are therefore best understood as strategic-level professional and expert assessments of stakeholder-generated content, rather than as community-weighted priorities.

Our findings advance implementation science theory in three ways. First, by integrating CFIR with participatory concept mapping, we demonstrate that stakeholder-perceived priorities do not distribute evenly across CFIR domains. Rather, stakeholders construct an implicit hierarchy where Inner Setting factors are viewed as both important and feasible, Outer Setting factors as important but less feasible, and Intervention Characteristics as less prominent overall. This empirically derived hierarchy challenges assumptions about equal domain weighting in implementation planning and suggests that CFIR-guided strategies should account for differential actionability across domains. Second, our results reveal that stakeholders’ mental models of AI implementation extend beyond CFIR’s traditional constructs. Participants generated nuanced themes around algorithmic transparency, data governance, and human-AI trust that suggest domain-specific expansions may be needed when applying CFIR to AI implementations. Third, this study demonstrates methodological value in combining systematic frameworks with participatory prioritization, showing that theory-driven identification paired with stakeholder-driven ranking yields more actionable guidance than either approach alone; future framework development should consider technology-specific extensions that address unique characteristics of AI and digital health tools while maintaining consistency with core implementation science principles.

Beyond the overall cluster-level pattern, the go-zone quadrants offer three informative observations. First, although governance-related statements were, in aggregate, rated lower than benefit-oriented statements, the single most important statement in the entire set was a governance statement: human oversight during early implementation. Together with the high placement of accountability and oversight of AI, this suggests the panel did not deprioritize governance wholesale, but rather distinguished proximal, locally actionable safeguards (oversight, end-user education, phased roll-out) from distal, system-level requirements (regulation, data ownership, corporate control of data) that individual organizations cannot resolve alone. Second, the high-importance/low-feasibility quadrant was dominated not by technical challenges but by acceptability and trust, patient and provider acceptability were among the highest-importance statements overall yet were rated among the least feasible to address, suggesting that the panel viewed social license, rather than technology, as the binding constraint on AI adoption in cancer care. Third, the placement of nuanced human tasks, such as detecting distress conveyed through the unsaid elements of a trusting therapeutic relationship, in the important-but-infeasible zone signals a perceived boundary on what AI should be expected to do, consistent with the strong rating for preserving clinician time for tasks requiring human expertise. Practically, this suggests deploying documentation, summarization, transcription, and triage supports first, paired with visible human-oversight and end-user education, and treating patient and provider acceptability as a measured outcome, not an assumption.

The consistent pattern of higher importance and feasibility ratings for benefit-oriented domains, particularly those related to workflow efficiency and clinical quality, suggests that stakeholders differentiate between AI applications that extend existing clinical practices and those requiring broader organizational or governance change, aligning with implementation science literature on the influence of workflow compatibility, perceived usefulness, and organizational readiness on early adoption[50,51].

The subcluster structure reveals how stakeholders distinguish between different types of implementation challenges and opportunities, aligning with CFIR’s domain architecture. Within challenges and safeguards (Cluster 1), stakeholders differentiated between implementation process concerns (Subcluster 1.1: oversight, training, phased rollout), inner setting ethical considerations (Subcluster 1.2: trust, validation, bias), outer setting regulatory issues (Subcluster 1.3: privacy, liability, governance), and intervention characteristics quality requirements (Subcluster 1.4: data quality, validation standards). Within the benefits cluster (Cluster 2), subclusters differentiated between research and knowledge applications (Subcluster 2.1), direct workflow improvements (Subcluster 2.2), clinical quality enhancements (Subcluster 2.3), and patient-facing communication tools (Subcluster 2.4). Higher-feasibility subclusters coalesced around tools integrating directly into existing clinical workflows (reflecting CFIR’s “complexity” and “compatibility” constructs) and alleviating documentation or triage burdens, most evident in Subclusters 2.2 and 2.4[52]. Conversely, less-feasible subclusters were dominated by system-level requirements such as data governance, algorithmic transparency, and regulatory clarity, concentrated in Subclusters 1.2, 1.3, and 1.4, which typically require organizational- or policy-level infrastructure rather than individual clinician action. Taken together, this contrast suggests near-term implementation efforts may benefit from emphasizing workflow-aligned tools while simultaneously building longer-term governance and system readiness.

Workflow support is viewed as an extension of existing practice norms, whereas governance-related functions are perceived as requiring broader infrastructural or cultural change, consistent with CFIR’s distinction between inner and outer setting constructs[53].

The equity-adjacent themes have been frequently highlighted for e-health technologies[54]. In line with those, an important caveat concerns the equity-adjacent themes identified in this study, including the digital divide, marginalization of minority groups, trust, and access (e.g., statements 10, 17, 67, and 93). Because individual statements could not be attributed to specific stakeholder groups, and because the rating panel was predominantly professional (with one patient-expert), we cannot determine whether these themes reflect the lived perspectives of patients and communities or professionals’ perceptions of those issues. The relatively low importance and feasibility ratings these statements received should be interpreted with particular caution in this light: a panel with greater patient and community representation might have prioritized them differently. Eliciting and weighting equity-related priorities directly from affected communities remains an important task for future participatory work.

These patterns have direct implications for sequencing implementation strategies, distinguishing workflow-embedded, early-stage targets from domains requiring longer-term, system-level investment; the go-zone analysis supports this sequencing logic by separating areas of perceived readiness from those requiring additional planning.

To operationalize this sequencing logic, quadrant placements suggest different classes of implementation strategies. For high-importance, high-feasibility items, the indicated step is piloting, e.g., ambient documentation or triage supports paired with co-located safeguards (designated oversight roles, end-user training, phased roll-out), corresponding to established ERIC strategies such as preparing champions, ongoing training, and audit and feedback[52]. For high-importance, low-feasibility barriers, “prioritization” means early, deliberate investment because they gate later scale-up: patient and provider acceptability, for instance, translates into involving patients and family members in governance structures and measuring acceptability as a formal implementation outcome. Low-rated system-level items (e.g., regulatory frameworks, data ownership) are largely beyond a single organization’s control, warranting policy monitoring rather than local remediation. These translations are illustrative: selecting and tailoring specific strategies remains a local, context-dependent exercise.

Taken together, this study offers a structured framework grounded in stakeholder-generated statements and prioritized by a multidisciplinary expert panel, assisting organizations in sequencing implementation activities. The thematic patterns show that AI implementation in cancer care is a layered challenge: even within broad categories, stakeholders distinguish between different types of applications, concerns, and requirements. By integrating cluster and subcluster analysis with a feasibility–importance prioritization framework, the findings provide a starting point for identifying specific areas where readiness is perceived versus domains likely to require additional investment, offering an initial structure that organizations may adapt and validate within their own local contexts.

### Limitations

While these findings provide insight into stakeholder perspectives on AI implementation in cancer care, several limitations should be considered. The expert panel (n = 13) was purposively sampled to ensure representation across key domains: clinical cancer care (n = 7), AI research and development (n = 4), health system administration (n = 1), and patient expertise (n = 1). This approach aligns with concept mapping methodology, which supports validity and reliability with relatively small samples, including single-digit participation in some cases[39], with diminishing gains beyond approximately 10 participants[52]. However, the limited sample size may affect the stability of finer-grained structures, particularly at the subcluster level. Furthermore, although the statement pool was generated by a diverse, multi-stakeholder group, the sorting and rating tasks were completed by a predominantly professional panel with only one patient-expert; the resulting priority structure therefore primarily reflects professional and expert perspectives, and the stakeholder-centered framing of this study applies most fully to the statement generation phase. We note that concept mapping seeks conceptual validity — a useful structuring of a domain by informed participants — rather than statistical representativeness of a population[39,41,42]; patient perspectives entered the project primarily through the statement generation phase. Nevertheless, limited patient involvement at the rating stage means the prioritization reflects predominantly professional judgments, and the resulting framework supports analytic rather than statistical generalization, with priority weightings warranting examination in future studies with broader patient and community participation. Subcluster distinctions should therefore be interpreted as indicative rather than definitive. The 48 workshop participants represented a range of professional roles, but the sample was geographically concentrated within Canada, primarily British Columbia, which may limit transferability to other healthcare systems and cultural contexts. The workshop-based recruitment approach may have introduced selection bias, as participants were likely already engaged with AI in cancer care and may not reflect the broader healthcare workforce. The absence of individual-level demographic data also prevents assessment of how well equity-relevant perspectives (e.g., those of patients from marginalized communities) were represented among statement contributors. Subgroup analyses (clinical n = 7, nonclinical n = 6) were limited by small numbers within each group, constraining power to detect differences. Relatedly, because all ratings were nested within 13 raters and statements are not independent observations, the reported *P* values should be interpreted cautiously as exploratory summaries of patterns within this panel rather than as formal inference to a broader population. Moreover, although raters were given a single action-oriented definition of feasibility spanning all statement types, feasibility may still have been construed somewhat differently for benefits, concerns, and priorities; residual heterogeneity in interpretation cannot be excluded, and go-zone placements should be read with this caveat in mind. The study also captured perspectives at a single point in time, and views may evolve as AI technologies change. Finally, the 2-stage clustering approach, while enabling nuanced interpretation, involved analytic decisions that may influence the resulting structure.

### Future Directions

Future research should extend this work to further inform AI implementation in cancer care. Future work could employ freelisting with salience analysis during statement generation to distinguish “top of mind” priorities from ideas mentioned but less central, grounding prioritization more directly in participant-generated data. Longitudinal, multi-institutional studies with larger and more diverse samples would help assess whether the cluster and subcluster structures remain stable over time and are transferable across cultural and organizational contexts. Empirical studies evaluating implementation strategies aligned with the identified priority domains, and testing the practical utility of this framework in real-world initiatives, would strengthen the evidence base and clarify how concept mapping outputs can support phased, context-sensitive AI adoption in cancer care.

## CONCLUSIONS

The integration of AI into cancer care holds substantial promise for improving clinical workflows and care delivery, but its successful adoption depends on how implementation challenges and opportunities are navigated in practice. Using concept mapping, this study provides a structured analysis of stakeholder-generated perspectives, prioritized by a multidisciplinary expert panel of how perceived benefits, concerns, and prerequisites for AI implementation relate to one another, offering insights beyond surface-level qualitative themes.

Our findings highlight a clear distinction between workflow-aligned AI applications that stakeholders view as both important and feasible, and system-level governance and accountability requirements that are perceived as highly important but less immediately actionable. This pattern underscores the value of a phased implementation approach, in which organizations pilot workflow-integrated tools that address immediate clinical pressures, paired with locally actionable safeguards such as human oversight and end-user training, while concurrently investing in longer-term governance, policy, and infrastructure development for the barriers that gate broader scale-up. By integrating cluster and subcluster analysis with feasibility–importance prioritization, this study offers an initial, context-specific framework that may support strategic planning and sequencing of AI implementation efforts in cancer care. These findings provide an initial foundation for future research and may assist healthcare organizations, implementation scientists, and policymakers in developing context-sensitive, sustainable approaches to AI adoption.

## ADDITIONAL FILES

1. **Additional file 1:** Supplemental Table 1. Statement composition of subclusters. Table presenting all 100 statements assigned to their respective subclusters (1.1-1.4 and 2.1-2.4), including statement IDs, full text, and mean importance and feasibility ratings. (.docx)
2. **Additional file 2:** Supplemental Table 2. Per-quadrant compilation of statements from go-zone analysis. Table presenting statements classified into four quadrants (High-High, Low-High, High-Low, Low-Low) based on their importance and feasibility ratings relative to the overall average, including statement IDs, full text, importance scores, and feasibility scores. (.docx)
3. **Additional File 3:** Supplemental Information. Standards for Reporting Implementation Studies (StaRI) — Populated Checklist. This study follows the Standards for Reporting Implementation Studies (StaRI)[55,56]; the populated checklist is provided as **Additional file 3**. (.docx)

## LIST OF ABBREVIATIONS

AI: Artificial intelligence
BC: British Columbia
CFIR: Consolidated Framework for Implementation Research
EB AI: Evidence–based artificial intelligence
MDS: Multidimensional scaling
UBC: University of British Columbia

## DECLARATIONS

### Ethics approval and consent to participate

This study was approved by the UBC Research Ethics Board (H24–03306). All workshop participants provided informed consent for data collection and research purposes.

### Consent for publication

Not applicable.

### Availability of data and materials

The dataset supporting the conclusions of this article are included within the article and its additional files. The code and scripts used for analysis are publicly available on GitHub (https://github.com/chhaviiiii/concept_mapping).

### Authors’ contributions

- Conceptualization: H.H.X., J.J.N., A.T.B., C.C., A.F.B.
- Methodology: H.H.X., J.J.N., A.T.B., C.C., A.F.B.
- Investigation: H.H.X., C.N., J.J.N., D.H., J.A., S.R., A.T.B., C.C., A.F.B.
- Data curation: C.N.
- Formal analysis: C.N.
- Software: C.N.
- Visualization: C.N.
- Writing – original draft: H.H.X., C.N.
- Writing – review & editing: All authors.
- Project administration: J.J.N.
- Supervision: J.J.N., A.F.B.
- All authors approved the final version of the manuscript.

## Acknowledgements

We thank all workshop participants at the 2024 BC Cancer Summit for their valuable contributions, as well as the expert panel members who completed the statement sorting and rating tasks for the concept mapping analysis.

