## Supplemental Table 1 for "Towards clinical implementation of artificial intelligence in cancer care: Concept mapping analysis of provincial workshop findings"

**Supplemental Table 1. Statement composition of subclusters**

| **Cluster** | **Subcluster** | **Statement ID** | **Statement** | **Importance** | **Feasibility** |
| --- | --- | --- | --- | --- | --- |
| 1 | 1.1 | 1 | Human oversight during implementation in early days | 4.25 | 3.42 |
|  | 1.1 | 16 | Proactive preventative care | 3.17 | 3.00 |
|  | 1.1 | 19 | Developing a rigorous privacy and quality assurance framework | 3.50 | 3.42 |
|  | 1.1 | 21 | Helps with generation of ideas can act as a sounding board | 2.83 | 3.58 |
|  | 1.1 | 29 | Data governance policies and regulations to enhance data quality and accountability | 2.92 | 2.58 |
|  | 1.1 | 32 | Regulation framework and a good understanding of the medico-legal implications | 2.83 | 2.25 |
|  | 1.1 | 37 | Consultation recording and transcription | 3.17 | 4.18 |
|  | 1.1 | 38 | Ensuring cybersecurity of AI tools | 3.75 | 3.17 |
|  | 1.1 | 39 | Educating end-users about properly navigating AI | 3.83 | 3.58 |
|  | 1.1 | 43 | Ensuring accessibility and translation | 3.50 | 3.67 |
|  | 1.1 | 45 | Embed research / quality improvement into all areas | 2.92 | 3.25 |
|  | 1.1 | 68 | Phased roll-out: Start with simple, "low stakes" tasks continue in stages | 3.17 | 4.08 |
|  | 1.1 | 88 | Ensuring interpretability of AI tools | 3.42 | 3.33 |
|  | 1.2 | 3 | Concerns with patient acceptability | 4.08 | 3.17 |
|  | 1.2 | 5 | False confidence | 3.33 | 2.73 |
|  | 1.2 | 7 | AI could perpetuate existing biases | 3.33 | 3.25 |
|  | 1.2 | 10 | Concerns with digital literacy and digital divide barriers | 3.08 | 2.75 |
|  | 1.2 | 20 | AI could be misused | 3.17 | 2.75 |
|  | 1.2 | 28 | Concerns about personalizing care | 2.83 | 3.25 |
|  | 1.2 | 33 | AI could lead to excessive human delegation, assuming that AI will take care of it! | 2.58 | 2.50 |
|  | 1.2 | 35 | AI could be of concern regarding its ongoing validation | 3.00 | 3.09 |
|  | 1.2 | 47 | Concerns with attrition of skills in healthcare providers | 2.58 | 2.58 |
|  | 1.2 | 49 | Loss of human connection/interactions | 3.33 | 2.75 |
|  | 1.2 | 52 | AI could engender issues with trust | 2.75 | 2.67 |
|  | 1.2 | 55 | AI hallucinations | 3.17 | 2.83 |
|  | 1.2 | 60 | Concerns with standards for quality, accuracy, etc. | 3.08 | 2.92 |
|  | 1.2 | 63 | Distress is nuanced and often detected in the unsaid of human communication and in a trusting therapeutic relationship | 3.42 | 2.50 |
|  | 1.2 | 65 | Concerns with difficulty to know older data for training and missing new breakthrough information | 2.92 | 2.92 |
|  | 1.2 | 67 | Racism bias through research (history of medical research) | 3.08 | 2.50 |
|  | 1.2 | 71 | Over reliance by physicians and reduced problem solving skills | 2.67 | 2.75 |
|  | 1.2 | 77 | Concerns with going too far and can't come back | 2.33 | 2.50 |
|  | 1.2 | 82 | AI could make mistakes | 3.33 | 3.00 |
|  | 1.2 | 83 | Concerns with stability of the system (during virus attack, power outage, system interruption) | 2.75 | 2.42 |
|  | 1.2 | 87 | Can existing healthcare servers support the use of AI? | 2.17 | 3.08 |
|  | 1.2 | 93 | Minority groups being marginalized | 3.00 | 2.92 |
|  | 1.3 | 2 | Concerns with confidentiality | 3.75 | 3.25 |
|  | 1.3 | 13 | Too much bureaucracy slowing progress | 3.00 | 3.00 |
|  | 1.3 | 14 | Concerns with liability | 3.33 | 2.83 |
|  | 1.3 | 26 | Concerns with job security | 2.42 | 2.25 |
|  | 1.3 | 36 | AI could be of concern to patient privacy | 3.08 | 3.08 |
|  | 1.3 | 40 | Concerns with protection of personal health identifiers | 3.42 | 3.00 |
|  | 1.3 | 54 | Concerns with responsibility accountability of AI recommendations | 3.17 | 3.00 |
|  | 1.3 | 61 | Concerns with power outages and downtime procedures | 2.17 | 2.58 |
|  | 1.3 | 66 | Concerns with consent of patient | 3.25 | 3.17 |
|  | 1.3 | 74 | Concerns with maintaining up to date information, sources and programming | 2.83 | 3.42 |
|  | 1.3 | 75 | Data privacy concerns | 3.33 | 2.83 |
|  | 1.3 | 78 | Control of data by corporations | 2.75 | 2.50 |
|  | 1.3 | 81 | Concerns with transparency | 3.08 | 2.75 |
|  | 1.3 | 86 | Lack of training/knowledge for those using the AI tool in the real world | 3.17 | 3.25 |
|  | 1.3 | 91 | Concerns that data is held by private companies | 2.92 | 2.42 |
|  | 1.3 | 95 | Data ownership and standards and guidelines on AI developed are not established | 2.83 | 2.92 |
|  | 1.3 | 96 | Possibility of data breaches / data leaks | 3.00 | 2.83 |
|  | 1.3 | 99 | Cybersecurity concerns | 2.75 | 2.42 |
|  | 1.4 | 9 | Ensuring accountability and oversight of AI | 4.00 | 3.58 |
|  | 1.4 | 12 | Diversification of AI training data and sources | 3.33 | 3.33 |
|  | 1.4 | 25 | Establishing proper consent | 3.00 | 3.08 |
|  | 1.4 | 46 | "Garbage in, garbage out" - data that the AI model is trained on must be good, clean, large and diverse | 3.58 | 2.83 |
|  | 1.4 | 48 | Preservation of human touch | 3.08 | 2.83 |
|  | 1.4 | 56 | Multidisciplinary collaboration and training in developing AI models | 3.17 | 3.25 |
|  | 1.4 | 73 | Ensuring clinical validation of AI tools | 3.50 | 3.42 |
|  | 1.4 | 85 | Establishing guidelines for "best practices" for training AI + clinical validation | 3.50 | 3.17 |
|  | 1.4 | 94 | Patient/parent/caregiver/provider acceptability | 3.92 | 2.92 |
| 2 | 2.1 | 6 | AI could provide alternate perspectives | 3.17 | 3.17 |
|  | 2.1 | 11 | Better detection (of moles, cancer lumps) | 3.58 | 3.75 |
|  | 2.1 | 15 | AI could decrease false positives | 3.42 | 3.42 |
|  | 2.1 | 24 | Benefits in early distress screening | 3.33 | 3.25 |
|  | 2.1 | 27 | Benefits with patient access to accurate information | 3.25 | 3.55 |
|  | 2.1 | 34 | Knowledge repositing for learning and research | 2.92 | 3.50 |
|  | 2.1 | 41 | Facilitation of clinical research | 3.08 | 3.42 |
|  | 2.1 | 42 | Improve access to services | 3.67 | 3.58 |
|  | 2.1 | 44 | AI could help improve accuracy | 3.58 | 3.75 |
|  | 2.1 | 51 | Collate large databases | 3.25 | 4.17 |
|  | 2.1 | 58 | Noticing patterns and new things humans haven't noticed | 3.58 | 3.50 |
|  | 2.1 | 62 | Increased opportunities for innovation | 2.92 | 3.58 |
|  | 2.1 | 97 | Potential for widening differential diagnosis, widening perspective of patient | 3.00 | 3.33 |
|  | 2.2 | 4 | Efficiency improved for documentation | 3.58 | 4.17 |
|  | 2.2 | 18 | AI could free up time for pt care | 3.75 | 3.67 |
|  | 2.2 | 23 | AI allows access vast information very quickly to help diagnosis | 3.75 | 4.00 |
|  | 2.2 | 30 | Savings in time, can integrate different information and improve in treatment especially when there is less number of human specialist | 3.83 | 3.67 |
|  | 2.2 | 31 | AI could remove human bias | 3.17 | 2.75 |
|  | 2.2 | 57 | 24/7 availability, AI doesn't get tired or brain fog | 3.42 | 4.25 |
|  | 2.2 | 59 | Prediction/prognosis/treatment recommendations | 3.92 | 3.58 |
|  | 2.2 | 70 | Potential in navigation and directing to self-management resources | 3.42 | 3.67 |
|  | 2.2 | 72 | Ability to make text/conversation more digestible for patients (less technical) | 3.50 | 3.83 |
|  | 2.2 | 76 | Triage diagnostic results for clinicians | 3.67 | 3.67 |
|  | 2.2 | 79 | AI could help address the human health resources issue | 3.58 | 3.25 |
|  | 2.2 | 80 | Faster assessments to inform decision-making | 3.50 | 3.58 |
|  | 2.2 | 84 | Live translation and/or transcription of conversations | 3.73 | 4.08 |
|  | 2.2 | 89 | Allow clinicians to focus on tasks requiring their expertise | 4.17 | 4.00 |
|  | 2.2 | 98 | Lists possible outcome and risks for the patient | 3.83 | 3.58 |
|  | 2.2 | 100 | Automation of routine tasks | 4.00 | 4.08 |
|  | 2.3 | 8 | Minimizing bias and increasing performance and accuracy | 4.00 | 3.33 |
|  | 2.3 | 17 | Benefits in equitable access | 3.33 | 2.92 |
|  | 2.3 | 53 | Pooling large volume of data can improve accuracy of AI system and allow for more timely diagnosis and treatment recommendation | 3.58 | 3.75 |
|  | 2.3 | 64 | Personalization of AI support tools for patients | 3.58 | 3.33 |
|  | 2.3 | 69 | Reduce admin burden | 3.92 | 4.17 |
|  | 2.3 | 90 | Identifying and focusing on high-priority areas, such as prevention | 3.08 | 3.42 |
|  | 2.4 | 22 | Benefits with accessibility & communication (translating languages lay terms) | 3.25 | 3.58 |
|  | 2.4 | 50 | Summarization of large amounts of text or data – increases digestibility/highlighting key points. | 3.67 | 4.33 |
|  | 2.4 | 92 | Maintaining competency with changing best practices and approved standards | 3.17 | 2.83 |
